# Surgical Information Assistant: an agentic information retrieval system for surgical information and a benchmark dataset

**DOI:** 10.1101/2025.05.20.25328046

**Authors:** Kiran Bhattacharyya

## Abstract

We present the Surgical Information Assistant, an agentic retrieval-augmented generation (RAG) system designed to improve access to surgical knowledge in resource-constrained settings. Built on the Open Manual of Surgery for Resource-Limited Settings, the assistant uses a retrieval-method we call DeRetSyn (Decom-pose–Retrieve–Synthesize). We evaluate DeRetSyn using automated metrics and partial human validation across 14,500 synthesized question–answer pairs and find that it achieves 63% top-1 accuracy using a 3B Llama model – outperforming GPT-4o (42.5%) without RAG and a 8B Llama model with conventional RAG (≃53%) while being significantly smaller and more computationally efficient. We also find that the DeRetSyn system with the Llama 3B model outperforms GPT-4o on the publicly available PubMedQA dataset on overall accuracy under specific prompting patterns. The Surgical Information Assistant demonstrates how agentic orchestration can extend the capabilities of small language models and offers a deployable framework for point-of-care medical decision support, education, and QA in low-bandwidth environments. We plan to release our benchmark dataset, codebase, prompt library, and RAG evaluation results for all categories for the entire dataset along with chain-of-thought reasoning from GPT-4o, Llama-3.1-8B, and Llama-3.2-3B upon publication.

## 1 Introduction

Large language models (LLMs) are increasingly being explored for medical question-answering (QA), offering the potential to democratize access to clinical knowledge and support decision-making in real time. However, in high-stakes fields like surgery—especially in low-resource settings— hallucinations, opaque reasoning, and retrieval failures can limit the effectiveness of even the most powerful general-purpose models. As evidence of these limitations, there is increasing interest in the community to benchmark these language models for specific tasks under different orchestrations and prompting patterns [1, 2, 3].

Retrieval-Augmented Generation (RAG) has emerged as a promising solution to mitigate these issues by grounding LLM outputs in retrieved, external knowledge [4, 5]. Notable examples include Almanac [6], which augments OpenAI’s text-davinci-003 with curated medical sources such as UpToDate and PubMed to improve factual accuracy (+18% over ChatGPT); and i-MedRAG [7], which iteratively prompts LLMs to reformulate complex medical queries, significantly outperforming vanilla RAG on medical licensing exam datasets such as MedQA and MMLU. The MedRAG Toolkit [7] enables systematic benchmarking across corpora (e.g., StatPearls, Merck Manual) and LLMs 39th Conference on Neural Information Processing Systems (NeurIPS 2025). (e.g., GPT-3.5, Mixtral-8x7B), showing that small models with domain-specific retrieval can rival GPT-4.

**Figure 1:**
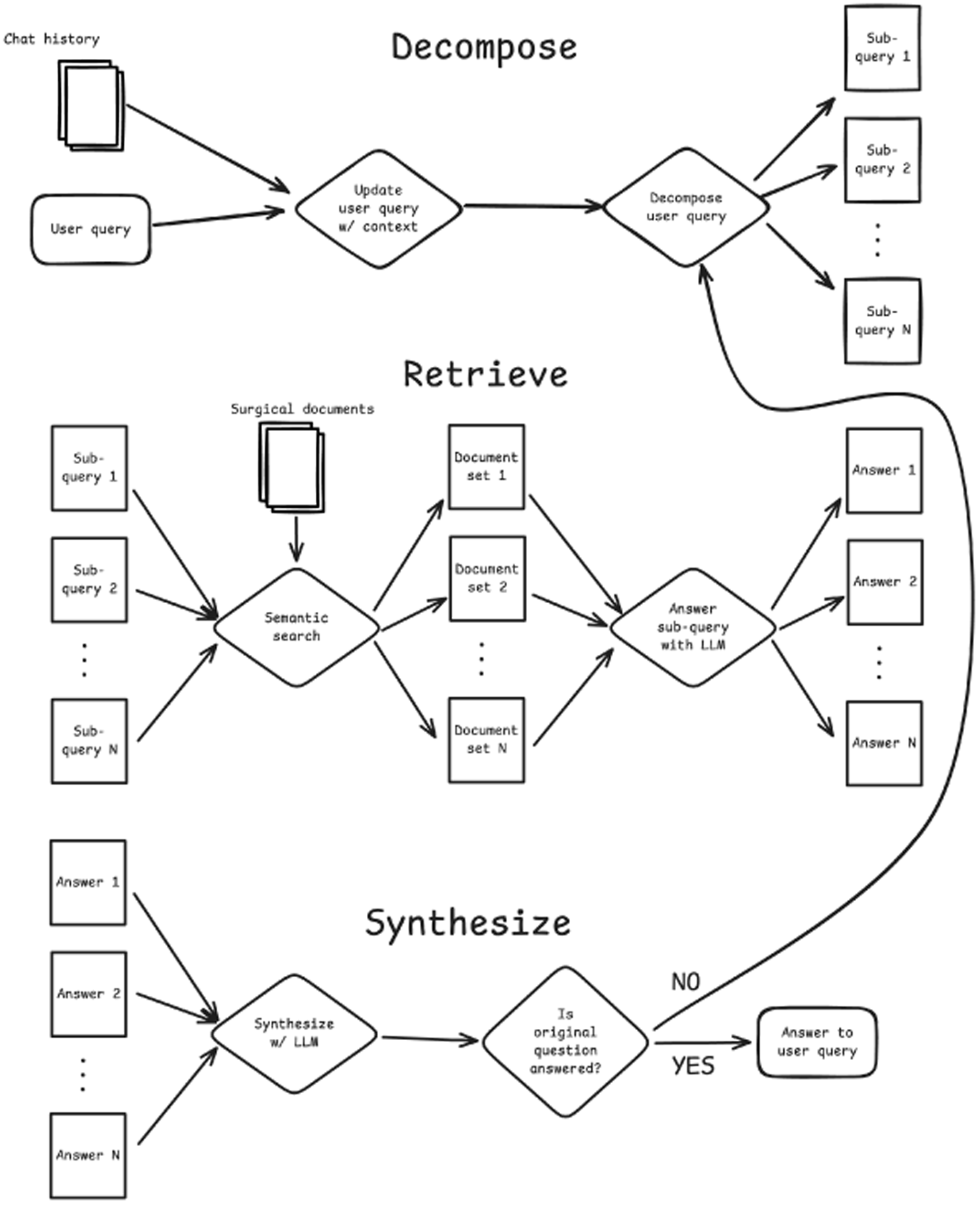
System design of the DeRetSyn pipeline.

Other advanced systems include PaperQA [8], which decomposes RAG into tool-like primitives (search, gather, synthesize) orchestrated by an agent and CRP-RAG [9], which uses reasoning graphs to plan multi-step retrieval. These approaches share a common goal: to go beyond naive single-step retrieval and embrace structured, iterative, and interpretable information-seeking workflows [10].

Despite these advances, most systems still use larger LLMs that cannot run on smaller devices, require stable internet access, and high resource availability—assumptions that do not hold in many practical healthcare settings. In such contexts, clinicians and educators require lightweight, interpretable, and reliable QA tools that can function offline while maintaining high domain accuracy.

In this paper, we introduce a Surgical Information Assistant that combines agentic RAG with deployment-focused design. Built on the Open Manual of Surgery for Resource-Limited Settings, the system enables LLM-based answers grounded in globally accessible, non-specialist surgical guidance. The system is powered by a modular agentic architecture we call DeRetSyn (Decom-pose–Retrieve–Synthesize), which— 1) decomposes complex surgical queries into sub-questions; 2) retrieves relevant text from a FAISS-indexed corpus; 3) synthesizes answers using a small Llama-3.2-3B model; 4) refines with iteration; and finally 5) falls back to separate document store or API service if corpus coverage is insufficient.

Inspired by agentic frameworks such as PaperQA and CRP-RAG, DeRetSyn orchestrates multi-agent workflows to emulate complex clinical reasoning. In a benchmark evaluation of 14,500 QA pairs, DeRetSyn outperforms GPT-4o (42.5% accuracy) and standard RAG with a larger Llama-3.1-8B model (35.8%), achieving 63.0% accuracy with a 3B model. The system also demonstrates competitive performance in the PubMedQA dataset across various configurations [11].

While the project primarily uses the Open Manual of Surgery in Resource-limited Settings (OMSRS) as its source documentation, this project is not officially associated with or endorsed by the OMSRS. The system is designed to build on OMSRS within the scope of the Creative Commons licensing structure. All code, data, prompts, evaluation results, and LLM outputs used for this manuscript will be made available upon publication in addition to hosting a functional application in the Streamlit community cloud.

Our core contributions are threefold:

- A deployable, reproducible agentic RAG system for surgical QA in resource-limited settings.
- A demonstration that agentic orchestration enhances small model performance in domain-specific retrieval.
- A publicly available benchmark, corpus, codebase, and prompt set to enable future research and reproducibility in clinical QA.

## 2 Methods

### 2.1 System architecture

This section outlines the key components and processes underlying the assistant’s functionality, emphasizing its agentic retrieval system, asynchronous and sequential processing capabilities, and fallback mechanism.

#### DeRetSyn: an agentic retrieval system

The core of the system is an agentic retrieval system, which employs a multi-step process to provide answers to questions [12, 13, 8]:

1. **Decomposition**: The user’s query is analyzed and processed to extract sub-questions.
2. **Retrieval**: Each sub-question is used to search a pre-built FAISS (Facebook AI Similarity Search) index containing vector representations of the surgical manual content.
3. **Synthesis**: An LLM attempts to answer the original question based on the sub-questions and their answers.
4. **Iteration**: An agent performs a check to determine if a satisfactory answer is generated for the original question. If a satisfactory answer is not generated, the system creates new sub-questions and repeats the process accumulating sub-queries and their answers over time [14].
5. **Best Effort**: After three iterations, if no satisfactory answer is found, the system is referred to a more general fallback data store as a secondary source. In this case, we use an openly available Wikipedia index (up-to-date until 2017) made available by the Dspy project [15] to provide the most general source of fallback information possible to handle potential out-of-domain queries.

The agentic retrieval system developed here is named DeRetSyn (after Decompose, Retrieve, and Synthesize).

#### Asynchronous and sequential processing

The DeRetSyn is designed to run parts of the retrieval process asynchronously, while other parts are executed sequentially. This hybrid approach allows for efficient processing of multiple sub-questions and retrieval tasks synchronously while maintaining the logical flow of the question-answering process with checks and iterations.

#### Fallback mechanism

When the primary surgical database doesn’t yield satisfactory results, the assistant employs a dual-search approach using using a fallback datastore [16]— 1) *Fast Search:* A rapid search is performed using ColBERTv2 on a pre-indexed Wikipedia abstract dataset which is publicly available [15]; 2) *Slow Search:* If a satisfactory answer is not found through Fast Search. A more comprehensive search is conducted using the Wikipedia Python library to retrieve full page content using an API service (requiring an internet connection); and 3) *Integration:* Results from both searches are combined with information from primary surgical texts to provide a comprehensive answer. Since this fallback system increases the memory footprint of the system, we perform ablation experiments where this mechanism is removed.

### 2.2 Implementation details

#### Core Components

The Surgical Information Assistant is implemented in Python and consists of several key components—1) a FAISS index; 2) LLMs; 3) asynchronous processing of certain tasks; 4) an API interface for programmatic use; and 5) a Streamlit interface for user-friendly web-based interactions.

#### Text embedding

The embedding was done by parsing the PDF documents from the Open Manual of Surgery in Resource-limited Settings to text. Then chunking the text into 1000 character elements with 200 character overlap between chunks. These character chunks were embedded into vectors with the all-MiniLM-L2-v2 model and indexed in a FAISS index with a meta-data file storing the file name and line-numbers for each embedding.

#### QA Dataset Generation

To validate DeRetSyn, a QA dataset was created using the following steps— 1) *Text Extraction*: PDFs from the surgical manual are processed to extract text content; 2) *Chunking*: The extracted text was divided into chunks of 500, 1000, 2000, 3000, 4000, or 5000 characters in length selected psuedo-randomly with equal probability; and 3) *QA Pair Generation*: A LLM was used to generate question-answer pairs from each fragment [17].

#### Model parameters and hardware specifications

We selected the Llama-3.2-3B model for our evaluations due to its strong performance on both medical and general-purpose benchmarks [18, 19]. As a member of the Llama family, it also enables a direct comparison with the larger Llama 3.1 8B model, allowing us to analyze how performance varies with model scale with different RAG paradigms and parametric knowledge.

The OpenAI API was used to call GPT-4o (March-August 2025). The TogetherAI API was used to evaluate all other models (March-August 2025). All inference was made with temperature=0 and all results are from top-1 responses.

## 3 Evaluation

### 3.1 Dataset Creation and Curation

To rigorously evaluate our surgical information retrieval system, we first created a comprehensive question-answer (QA) dataset derived from the Open Manual of Surgery in Resource-Limited Settings. We refer to this as the *OpenManualOfSurgeryQA dataset*. The dataset creation process involved:

- **Question Generation**: We programmatically generated over 16,000 question-answer pairs during the document embedding process, covering a wide range of surgical topics, procedures, and techniques contained within the source documents using Mistral Small 3.1 24B. The context used to generate the question-answer pairs were retained for quality review. Further details are provided below.
- **Quality Review**: The generated question-answer pairs underwent both automated filtering and manual review to ensure relevance, clarity, and faithfulness to the original context from which they were generated. Questions that were ambiguous or poorly formulated were eliminated.
- **Dataset Refinement**: After the quality review process, the dataset was reduced to 14,529 high-quality question-answer pairs, which served as our evaluation benchmark. Note that all question-answer pairs were not reviewed manually due to resource limitations. Details about quality review are provided below.

#### Question-answer generation prompt

The prompt consists of several critical instructions. First, it mandates strict *groundedness*, requiring that answers be derived exclusively from the provided text. Second, it enforces *self-containment*, ensuring questions are fully understandable without the source passage. Third, it directs the model to create questions that are *research-able* having enough context that relevant information and passages can be retrieved to answer the question. Finally, the prompt guides the model to generate a diversity of question types (e.g., definition, causality, comparison, yes/no) and to synthesize answers concisely rather than merely extracting sentences verbatim. Lastly, the prompt mandates a step-by-step reasoning process and a structured output format. We also include a 1-shot example to help encourage instruction-following and format consistency.

#### Quality review process for QA-pairs

The manual review process included inspecting 1000 generated QA pairs along with the context used to generate them to identify ambiguous questions and the patterns within them. This was done by 2 individuals, one of whom is a practicing clinician. The patterns found in ambiguous questions were—1) direct references to the text fragment used to generate the QA pair with terms such as “this passage”, “the passage”, “this text” or “the text”; 2) references to images within the documents that were associated to the text fragment with terms like “what is shown”, “the figure”, or “the image”; and 3) indirect or assumed references to information in other parts of the source document. All QA pairs where the question had any of the previous phrases were identified with sub-string search and removed. Other qualities of questions and answers that made them unsuitable were more difficult to remove with only sub-string search. Therefore, each remaining QA pair was further validated by an LLM (Mistral Small 3.1 24B) with reference to the passage used to generate it. The LLM was asked to confirm or deny each of the following criteria as they apply to the question or answer— 1) The question is answerable using the information in the passage; 2) The question is understandable completely without needing direct reference to the passage; 3) The question is stated with adequate clarity and specificity so that someone could search the internet to find the answer; 4) The answer must completely address all aspects of the question; 5) All informational content in the answer must be present in the passage; and 6) The answer must not assume context that is not present in the question.

If any of the previous criteria were denied, the QA pair was discarded. After this filtering through an LLM, an additional 1000 randomly selected remaining QA pairs were again manually reviewed. No ambiguous questions were found during this second round of review.

### 3.2 Evaluation Methodology

We conducted a series of comparative evaluations to assess the performance of various retrieval-augmented generation (RAG) strategies, as well as the baseline capabilities of language models without retrieval support. Our evaluation framework was guided by three primary research questions:

1) Can smaller language models augmented with external knowledge sources achieve accuracy comparable to or exceeding that of larger models operating without retrieval? 2) How do different RAG paradigms perform when applied to complex, domain-specific surgical question-answering tasks? [20, 21, 22] 3) How does DeRetSyn perform in comparison, with and without its core indexed information and the fallback?

#### LLM-as-judge

For each system configuration, we measured accuracy by comparing the generated responses to ground-truth answers. We used a LLM-as-judge (Mistral Small 3.1 24B) to determine whether each generated response matched the expected answer. The LLM was directed to use the following criteria. The generated answer must be

- *Factually consistent:* not contradict any part of the ground truth answer.
- *Complete:* address all parts of the question and include all critical information present in the ground truth answer.
- *Relevant:* must not present information that is irrelevant to the question The LLM-as-judge was also provided 3 few-shot examples in the prompt.

#### Human validation of LLM-as-judge

We performed a human evaluation for 100 generated responses to validate the LLM-as-judge. We psuedo-randomly selected 50 responses where the LLM-as-judge determined that the generated response was incorrect and 50 where it determined the response was correct. The human used the same criteria as listed above but was blinded to the decision of LLM-as-judge. We found a Cohen’s Kappa score of 0.9 between human and LLM-as-judge evaluations suggesting a high degree of agreement.

#### Ablation and Comparison Studies

To evaluate the contribution of key components in the DeRetSyn architecture, we conducted ablation and comparison studies across two datasets: PubMedQA [11] and our own OpenManualOfSurgeryQA benchmark.

On PubMedQA, we compared DeRetSyn against Chain-of-Thought (CoT) prompting [23] under two conditions: (1) when the model relied solely on parametric knowledge with no context provided, and (2) when it was given a fixed, relevant context for each question. Furthermore, to isolate the effects of agentic orchestration from retrieval, we ablated DeRetSyn’s retrieval and fallback modules, forcing it to answer either with no context or with predefined retrieval results—the context for each PubMedQA question. In the no-retrieval and no-context setting, the sub-queries in the DeRetSyn process were answered by model’s parametric knowledge. These comparisons allowed us to assess whether DeRetSyn improves over CoT both in zero-context settings and when relevant evidence is accessible. Finally, we evaluated the performance of the complete DeRetSyn system including retrieval from the OMSRS index and Wikipedia fallback on the PubMedQA dataset but without the provided context for each PubMedQA question. For all evaluations completed with the PubMedQA dataset, we use the “reasoning required” configuration where the reasoning is never provided for any question and we use the “pqa_labeled” subset of the data.

On the OpenManualOfSurgeryQA dataset, we extended these comparisons to include the ReAct prompting framework [24] and a vanilla RAG pipeline, evaluated across multiple model scales with the same OMSRS document index. We also performed two ablations: disabling the fallback mechanism to assess performance when broader-domain sources like Wikipedia are unavailable, and removing the decomposition module to simulate shallow single-step query handling. These studies help clarify the specific impact of decomposition, retrieval, and fallback on overall system performance and robustness.

## 4 Results

### Average response times and iteration counts

The average time to the final answer for the DeRetSyn system with the Llama-3.2-3B model when using the Together AI API was 14.2 seconds. The same measure on a M2 Macbook Pro used for local experiments was 8.2 seconds but multiple questions could not be processed synchronously. The average number of iterations for the DeRetSyn system was 1.9 across all experiments.

### Evaluations on PubMedQA

Critically, providing relevant context improves both CoT prompting and DeRetSyn substantially (Table 2). DeRetSyn with fixed PubMedQA context and no fallback achieves the highest accuracy (70.1%), outperforming CoT with the same context (63.5%) and GPT-4o under similar conditions (64.1%). Notably, DeRetSyn demonstrates robust gains even when retrieval and fallback are ablated: with only its agentic sub-querying pipeline and no access to external context, it still outperforms CoT without context (52.1% vs. 50.0%). When using its full architecture—including retrieval from the Open Manual of Surgery (OMSRS) and fallback but no access to the PubMedQA-provided context—DeRetSyn achieves 61.5% accuracy, approaching performance levels attained with explicit PubMedQA context. These results highlight the benefit of DeRetSyn’s agentic decomposition and orchestration not only in maximizing context utility but also in improving reasoning under constraints.

**Table 1:**
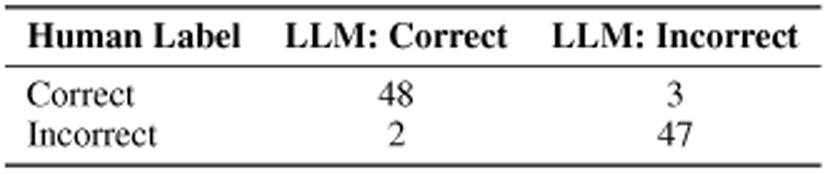
Confusion matrix comparing human evaluation and LLM-as-judge labels for 100 QA pairs.

| Human Label | LLM: Correct | LLM: Incorrect |
| --- | --- | --- |
| Correct | 48 | 3 |
| Incorrect | 2 | 47 |

**Table 2:**
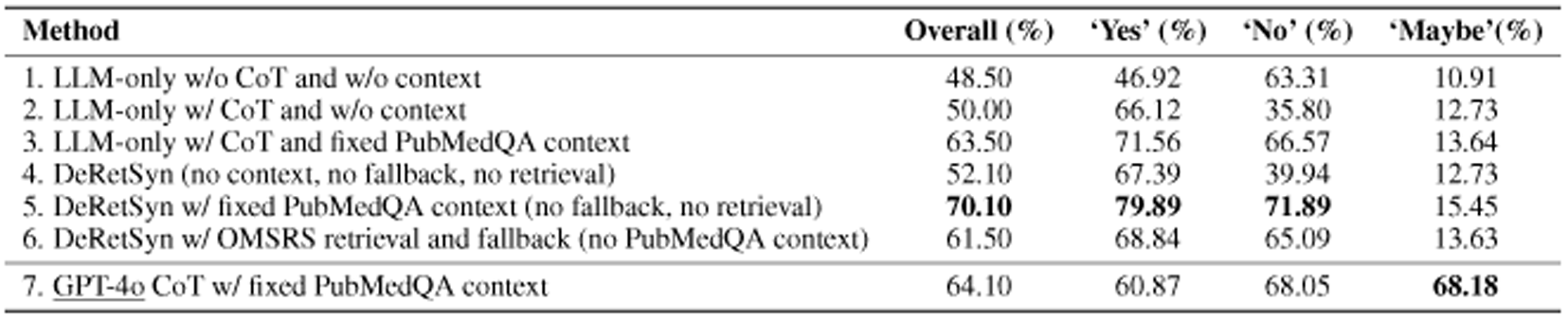
Performance on PubMedQA under different prompting and retrieval configurations. Accuracy is reported overall and per answer type for Llama-3.2-3B. The best performance for each category is bolded.

### Evaluation on OpenManualOfSurgeryQA

We evaluated eight distinct model configurations, including both non-augmented (base instruction-finetuned) models and several variants of retrieval-augmented generation (RAG) systems. All RAG-based models utilized a consistent FAISS index and identical context length settings to ensure comparability. We find that DeRetSyn with fallback performs the best overall (Table 3). The fallback to the secondary source occurred in ∼5% of cases.

**Table 3:**
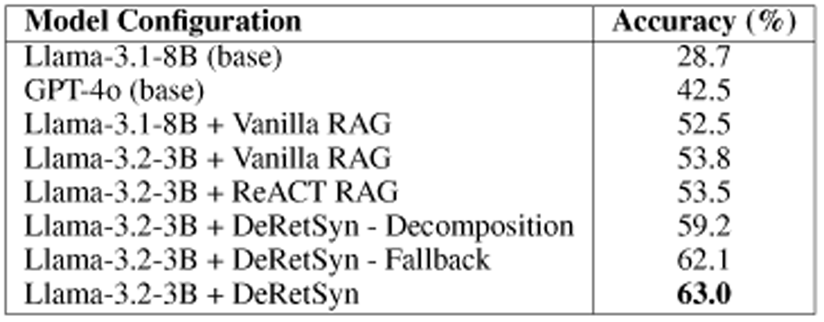
Accuracy comparison of different model configurations on the surgical information retrieval task for the OpenManualOfSurgeryQA (our) dataset.

To assess the variability and robustness of these results, we conducted a bootstrap analysis by sampling 30 subsets from the QA dataset, each consisting of 200 questions (Figure 2). This allowed us to estimate the standard deviation of each model’s accuracy, offering insight into performance stability and the likely user experience when only a limited number of queries are issued.

**Figure 2:**
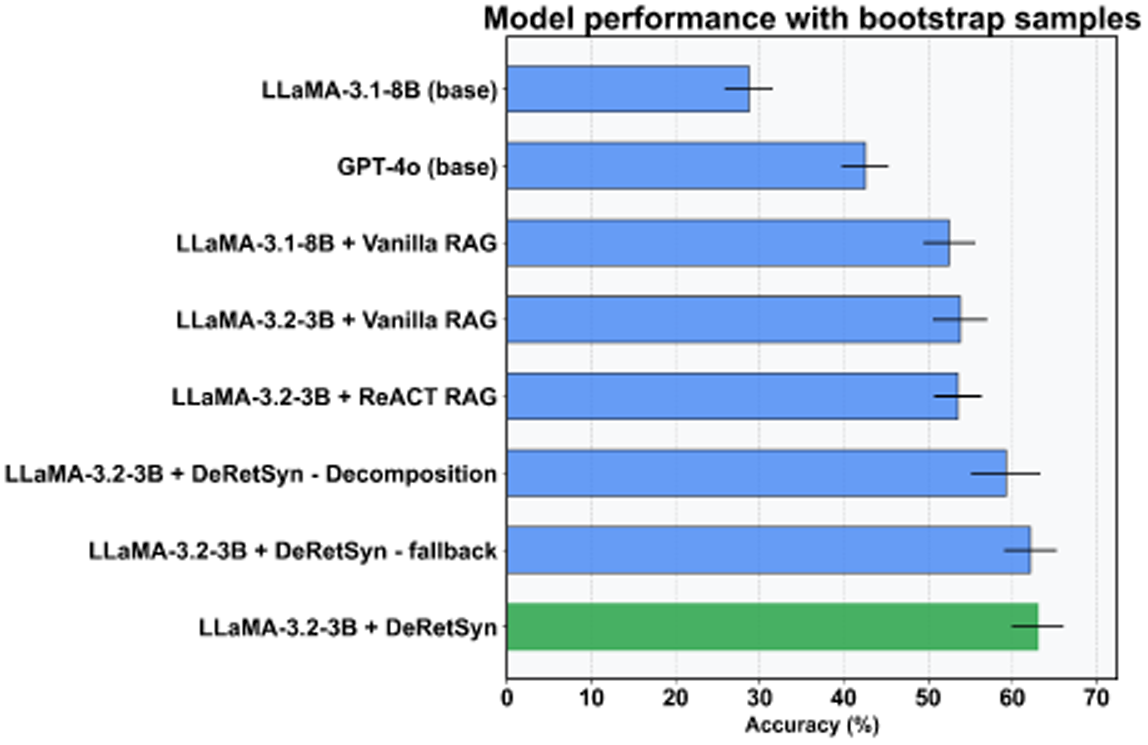
Accuracy and variability of model configurations based on bootstrapped evaluation samples. Error bars represent standard deviation across 30 samples of size 200.

We also computed the average number of tokens generated per response (Figure 3). Interestingly, Llama-3.2-3B generally used fewer tokens than the other LLMs tested in this study under similar configurations. However, the DeRetSyn system generates a larger number of total tokens than other RAG configurations due to the multiple generations required for each final response.

**Figure 3:**
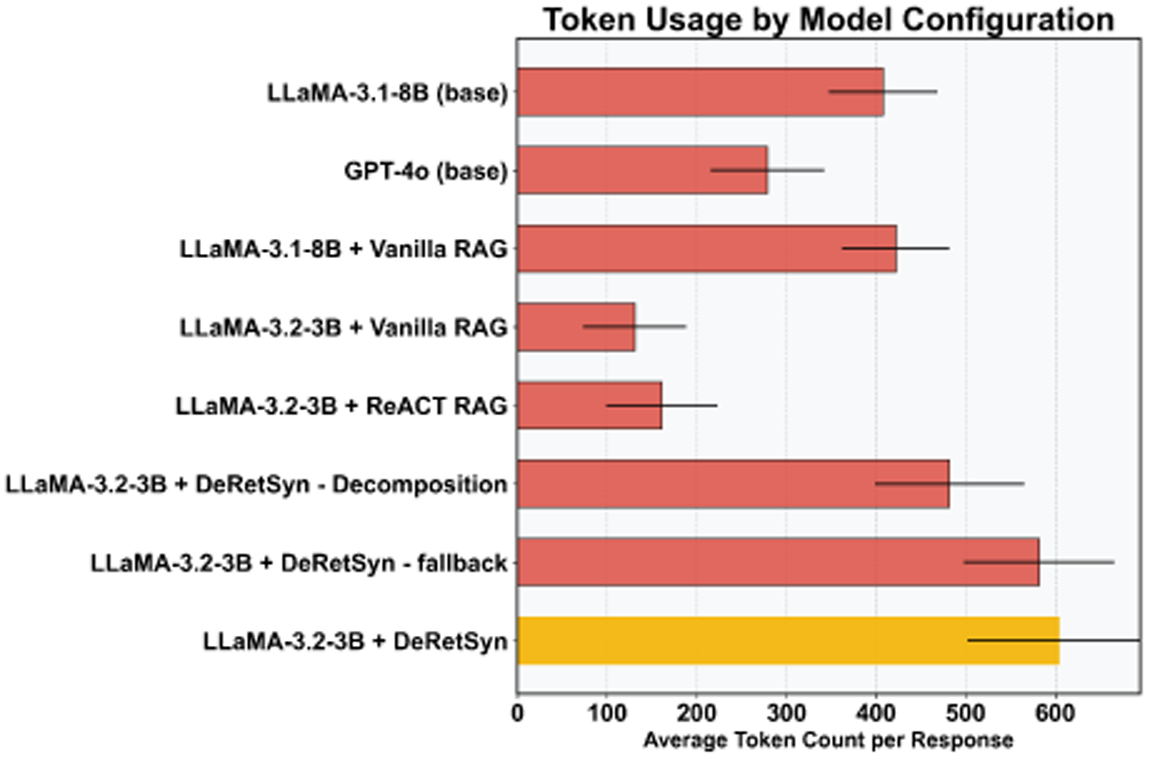
Number of tokens generated from each configuration evaluated. Error bars represent the standard deviation in the dataset.

### Observed Failure Modes

While DeRetSyn outperforms baseline methods, upon review with a practicing clinician, we observed several recurring failure patterns that inform directions for future improvement. *Sub-question misalignment* occasionally results from decompositions that are too broad, too narrow, or miss the clinical focus of the original query, leading to irrelevant or redundant retrievals. *Synthesis drift* occurs when retrieved content is sparse or overlapping, causing the model to blend accurate and fabricated details or repeat information without adding depth. *Fallback limitations* are evident when questions fall outside the domain of the indexed manual. In such cases, fallback to Wikipedia sometimes returns generic or lay summaries lacking clinical specificity. These failure modes suggest opportunities for improved decomposition quality control, fallback corpus curation, and tighter synthesis grounding in retrieved evidence.

## 5 Discussion

The evaluation results offer several key insights that we frame with comparisons:

1. **Parametric knowledge vs. RAG**: All RAG-augmented models substantially outperformed their non-augmented counterparts. For instance, augmenting Llama-3.1-8B with vanilla RAG improved its accuracy from 28.7% to 52.5%, underscoring the critical role of retrieval in enhancing performance (Table 3).
2. **Model Size vs. Knowledge Access**: The smaller Llama-3.2-3B model with vanilla RAG (53.8%) outperformed the much larger GPT-4o without retrieval (42.5%, Table 3). This supports our hypothesis that access to relevant external knowledge can allow smaller models to exceed the capabilities of larger models that rely solely on parametric memory [6, 25].
3. **DeRetSyn vs. CoT**: DeRetSyn outperforms CoT-prompting with relevant context (70.1% vs. 63.5%) and without (52.1 % vs. 50.0 %, Table 2). This suggests that DeRetSyn is better able to utilize parametric knowledge and provided context to generate an answer.
4. **DeRetSyn vs. other RAG Strategies**:The DeRetSyn method achieved the highest accuracy at 63.0%, outperforming both vanilla RAG and ReACT RAG on the same base model (Table 3). This result highlights the value of structured query decomposition, retrieval, and iterative synthesis.
5. **Performance vs. Efficiency Tradeoff**: While DeRetSyn provided superior accuracy, it also introduces additional latency and computational cost due to its multi-step architecture and increased number of tokens generated and language model calls.

Ablation experiments further elucidated the contributions of specific components. The fallback mechanism, which retrieves from a broader knowledge base (Wikipedia), had minimal impact on performance in this study. This likely reflects the fact that all evaluation questions were answerable using the core OMSRS dataset. In real-world deployments, where user questions may go beyond the primary domain, fallback retrieval may be more beneficial.

Removing the decomposition step reduced performance (from 63.0% to 59.2%) but still yielded results superior to other baselines. This is likely due to the model’s iterative refinement process, which prompts the generation of sub-questions when the system determines that answer is incomplete.

### Interpretability and Modularity of Agentic Orchestration

DeRetSyn’s modular agentic architecture offers interpretability, traceability, and reproducibility by surfacing intermediate steps—such as sub-questions, retrievals, and synthesized answers that can be inspected or rerun in isolation. In high-stakes domains like medicine, such visibility is essential for building trust and managing failure modes.

DeRetSyn mirrors principles from recent work on *self-consistency* [26], *universal self-consistency* [27], and *Tree-of-Thoughts* [28], which show that reasoning improves when multiple solution paths are generated, evaluated, and selectively refined. By decomposing complex queries into focused retrieval tasks and iteratively synthesizing answers, DeRetSyn effectively constructs a reasoning tree with embedded checkpoints. Its modularity not only enables reuse and correction of intermediate outputs without restarting the entire process but also supports extensibility—e.g., adding fallback layers, caching routines, or human-in-the-loop verification. This structure aligns with broader goals in interpretable AI, where reasoning provenance and fine-grained control are critical to safe deployment.

### Computational Considerations

The DeRetSyn pipeline delivers accuracy gains at the cost of greater computational complexity through a multi-stage process that includes query decomposition, parallel retrieval, and information synthesis. While each stage incurs additional token usage, the 9.2 percentage point improvement over vanilla RAG may justify these costs in clinical informatics applications. The Llama-3.2-3B implementation of DeRetSyn generated ∼5 times as many tokens as the configuration generating the least (Figure 3). However, this number could be reduced with specific prompting patterns [29].

The Llama-3.2-3B model generates 17–23 tokens/second on modern mobile devices as benchmarked by the developer community [30, 31, 32, 33]. Assuming an average of 600 tokens to arrive at the final response, inference time per query is estimated at ∼30 seconds. Devices capable of running multiple model instances in parallel may reduce this time through concurrent execution. Moreover, designed user interaction with the intermediate steps of the DeRetSyn system may be important for transparency and auditability (refer to Section 5) making the time-to-final response not the sole focus of usability.

### Limitations

Despite promising results, this study has several limitations. One of our evaluations relied on a synthetic QA dataset generated from the same corpus used for retrieval. While care was taken to avoid direct overlap (Section 2.2), this may still introduce evaluation circularity. However, DeRetSyn’s strong performance on PubMedQA helps support a claim of generalizability.

The QA generation process used heuristic prompts, which may bias the dataset toward fact-based questions and under-represent complex reasoning, ambiguity, or ethical judgment. However, this is not what we found from a manual review of a subset of questions, as described in Section 3.1.

Finally, we have not yet conducted prospective or interactive evaluations. Real-world testing with clinicians—especially in or simulating low-resource settings—is essential to assess hallucination risk, latency tolerance, and clinical integration in practice.

## 6 Conclusion

We show that small language models can deliver competitive performance on specialized tasks through advanced retrieval algorithms, such as the DeRetSyn system, making them suitable for resource-constrained environments [34, 14]. Additionally, DeRetSyn exemplifies the power of agentic retrieval systems in accessing and synthesizing critical surgical information while also providing transparency and auditability taking a step towards ensuring access to relevant medical information that also localize health data to reduce privacy concerns and connectivity dependence [35].

## Data Availability

All data and code is made available through a public Github repository.

https://github.com/MiningMyBusiness/surgical-information-assistant

## References

[1] Yu He Ke, Liyuan Jin, Kabilan Elangovan, Hairil Rizal Abdullah, Nan Liu, Alex Tiong Heng Sia, Chai Rick Soh, Joshua Yi Min Tung, Jasmine Chiat Ling Ong, Chang-Fu Kuo, et al. Retrieval augmented generation for 10 large language models and its generalizability in assessing medical fitness. npj Digital Medicine, 8(1):187, 2025.

2. Karen Ka Yan Ng, Izuki Matsuba, and Peter Chengming Zhang. Rag in health care: a novel framework for improving communication and decision-making by addressing llm limitations. NEJM AI, 2(1):AIra2400380, 2025.

[3] Yucheng Shi, Shaochen Xu, Tianze Yang, Zhengliang Liu, Tianming Liu, Xiang Li, and Ninghao Liu. Mkrag: Medical knowledge retrieval augmented generation for medical question answering. In AMIA Annual Symposium Proceedings, volume 2024, page 1011, 2025.

[4] Josip Vrdoljak, Zvonimir Boban, Marino Vilović, Marko Kumrić, and Joško Božić. A review of large language models in medical education, clinical decision support, and healthcare administration. In Healthcare, volume 13, page 603. MDPI, 2025.

[5] David J Bunnell, Mary J Bondy, Lucy M Fromtling, Emilie Ludeman, and Krishnaj Gourab. Bridging ai and healthcare: A scoping review of retrieval-augmented generation—ethics, bias, transparency, improvements, and applications. medRxiv, pages 2025–04, 2025.

[6] K Singhal and et al. Retrieval-augmented language models for clinical medicine. Nature, 620: 282–289, 2023.

[7] Guangzhi Xiong, Qiao Jin, Xiao Wang, Minjia Zhang, Zhiyong Lu, and Aidong Zhang. Improving retrieval-augmented generation in medicine with iterative follow-up questions. In Biocomputing 2025: Proceedings of the Pacific Symposium, pages 199–214. World Scientific, 2024.

[8] Hao Zhang and, et al. Discuss-rag: Enhancing retrieval-augmented generation via agent-led multi-turn reasoning for medical qa. arXiv preprint arXiv:2504.21252, 2024.

[9] Kehan Xu, Kun Zhang, Jingyuan Li, Wei Huang, and Yuanzhuo Wang. Crp-rag: A retrieval-augmented generation framework for supporting complex logical reasoning and knowledge planning. Electronics, 14(1):47, 2024.

[10] Siru Liu, Allison B McCoy, and Adam Wright. Improving large language model applications in biomedicine with retrieval-augmented generation: a systematic review, meta-analysis, and clinical development guidelines. Journal of the American Medical Informatics Association, page ocaf008, 2025.

[11] Qiao Jin, Bhuwan Dhingra, Zhengping Liu, William Cohen, and Xinghua Lu. PubMedQA: A Dataset for Biomedical Research Question Answering. In Proceedings of the 2019 Conference on Empirical Methods in Natural Language Processing and the 9th International Joint Conference on Natural Language Processing (EMNLP-IJCNLP), pages 2201–2211, 2019.

[12] Lu Chen and et al. Agentic retrieval-augmented generation: A survey. arXiv preprint arXiv:2501.09136, 2024.

[13] Jong Lee and et al. Surgraw: Structured multi-agent workflow with chain of thought reasoning for robotic-assisted surgery qa. arXiv preprint arXiv:2503.10265, 2024.

14. Ryan Ahsan. Clinical reasoning with rag: Building a local medical qa system, 2023. https://medium.com/@ahsan.new252/clinical-reasoning-with-rag.

[15] Omar Khattab, Arnav Singhvi, Paridhi Maheshwari, Zhiyuan Zhang, Keshav Santhanam, Sri Vardhamanan, Saiful Haq, Ashutosh Sharma, Thomas T Joshi, Hanna Moazam, et al. Dspy: Compiling declarative language model calls into self-improving pipelines. arXiv preprint arXiv:2310.03714, 2023.

16. Matillion. Agentic rag: Enhancing ai workflows, 2024. https://www.matillion.com/learn/blog/agentic-rag.

17. Saurav Joshi. Retrieval-augmented generation for medical qa with llama-2–7b, 2023. https://medium.com/@sauravjoshi23/retrieval-augmented-generation-for-medical-question-answering-with-llama-2-7b.

18. K Aaditya. Performance Comparison: Llama-3.2 vs. Llama-3.1 LLMs and Smaller Models (3B, 1B) in Medical and Healthcare AI Domains. Hugging Face Blog, Sep 2024. URL https://huggingface.co/blog/aaditya/llama3-in-medical-domain.

[19] NVIDIA. Llama-3.2-3b-instruct Model by Meta - NVIDIA NIM APIs. https://build.nvidia.com/meta/llama-3.2-3b-instruct/modelcard, 2024. Accessed: August 4, 2025.

[20] Yujia Wu and, et al. Mirage: Benchmarking retrieval-augmented generation for medicine. arXiv preprint arXiv:2402.13178, 2024.

21. A Liu and et al. Bias evaluation and mitigation in retrieval-augmented medical qa. arXiv preprint arXiv:2503.15454, 2024.

22. A Roberts and et al. Influence of context size and model choice in rag systems. In Findings of NAACL, 2025.

[23] Jason Wei, Xuezhi Wang, Dale Schuurmans, Maarten Bosma, Fei Xia, Ed Chi, Quoc V Le, Denny Zhou, et al. Chain-of-thought prompting elicits reasoning in large language models. Advances in neural information processing systems, 35:24824–24837, 2022.

[24] Shunyu Yao, Jeffrey Zhao, Dian Yu, Nan Du, Izhak Shafran, Karthik Narasimhan, and Yuan Cao. React: Synergizing reasoning and acting in language models. In International Conference on Learning Representations (ICLR), 2023.

[25] Z Wang and, et al. Medgraphrag: Safe medical llm via graph-based retrieval-augmented generation. arXiv preprint arXiv:2408.04187, 2024.

[26] Xuezhi Wang, Jason Wei, Dale Schuurmans, Quoc Le, Ed Chi, Sharan Narang, Aakanksha Chowdhery, and Denny Zhou. Self-consistency improves chain of thought reasoning in language models. arXiv preprint arXiv:2203.11171, 2022.

27. Xinyun Chen, Renat Aksitov, Uri Alon, Jie Ren, Kefan Xiao, Pengcheng Yin, Sushant Prakash, Charles Sutton, Xuezhi Wang, and Denny Zhou. Universal self-consistency for large language model generation. arXiv preprint arXiv:2311.17311, 2023.

[28] Shunyu Yao, Dian Yu, Jeffrey Zhao, Izhak Shafran, Thomas L Griffiths, Yuan Cao, and Karthik Narasimhan. Tree of thoughts: Deliberate problem solving with large language models, 2023. *URL* https://arxiv.org/abs/2305.10601, 3:1, 2023.

[29] Silei Xu, Wenhao Xie, Lingxiao Zhao, and Pengcheng He. Chain of draft: Thinking faster by writing less. arXiv preprint arXiv:2502.18600, 2025.

[30] Arm Newsroom. Ai inference everywhere with new llama llms on arm. https://newsroom.arm.com/news/ai-inference-everywhere-with-new-llama-llms-on-arm, 2024. URL https://newsroom.arm.com/news/ai-inference-everywhere-with-new-llama-llms-on-arm. Accessed: 2025-06-06.

31. Qualcomm. qualcomm/Llama-v3.2-3B-Instruct. Hugging Face, Sep 2024. URL https://huggingface.co/qualcomm/Llama-v3.2-3B-Instruct.

32. Hotellnx. Run Llama 3.2 3B on Phone - on iOS & Android. Reddit, Oct 2024. URL https://www.reddit.com/r/LocalLLaMA/comments/1fppt99/run_llama_32_3b_on_phone_on_ios_android/.

33. ctrl-brk. Phone LLM’s benchmarks? Reddit, Nov 2024. URL https://www.reddit.com/r/LocalLLaMA/comments/1glx6a5/phone_llms_benchmarks/.

[34] T Nguyen and et al. Systematic analysis of rag-based medical qa systems in low-resource settings. Information, 6(4):116, 2023.

[35] X Zhao and et al. Aipatient: Simulating patients with ehrs and agentic workflow. arXiv preprint arXiv:2409.18924, 2024.

